# THE EFFECT OF HAND HYGIENE INTERVENTION ON THE KNOWLEDGE AND SKILLS LEVEL OF SCHOOL CHILDREN

**DOI:** 10.1101/2022.01.15.22268666

**Authors:** Nurlela Mufida, Moses Glorino Rumambo Pandin

## Abstract

**Background:** Hand washing is an alternative, effective and inexpensive method that can be used to prevent infectious diseases. Washing hands with soap can improve children’s health. The purpose of this study was to determine the effect of hand hygiene intervention on the level of knowledge and skills of elementary school students. The method used to conduct this research is a critical review, namely by reviewing articles from databases such as Google Scholar, Pubmed, Direct, Medline. The search keywords used were: quantitative studies on delivery, intervention, hand hygiene, skills, children, and primary school. The selected articles are articles that meet the standard writing criteria, which were published between 2017-2021, both in English and Indonesian. Based on these evaluation criteria, 20 (twenty) studies were obtained. From the results of a literature search, several studies on the use of intervention strategies to provide hand hygiene health education found that although there were other intervention strategies to improve hand hygiene, there were significant differences in both knowledge and skills after being given the intervention.

## INTRODUCTION

School-age is an important period for the formation of clean and healthy living habits in children, who in the future are expected to become agents of change who can encourage healthy living in schools, communities, and the home environment (Nuraida, 2015). One of the strategic places in promoting health is Educational Institutions. Habits carried out by a child at school will also be carried out at home which is expected to influence the behavior of other family members (Solehati, 2015).

Hand hygiene is cleaning dirt by washing with water which can inhibit or kill bacteria acquired on the skin of the hands due to human contact with the environment, thereby preventing infections transmitted by hands. Failure to perform proper hand hygiene is a major cause of infection and the spread of resistant microorganisms (Hidayah & Ramadhani, 2019).

The habit of washing hands to maintain cleanliness is often forgotten and not noticed by many people, even though this habit has a positive influence that can affect the improvement of the health status of themselves and those around them.

The results of the literature reveal that school children have not realized that clean living behaviors such as the habit of washing hands in daily life, both in the school environment and in the playground, can improve their health. Children often eat food using unwashed hands. This habit is one of the factors that contribute to the incidence of diarrheal diseases and other infectious diseases. Based on the above phenomenon, the authors are interested in conducting a literature search as a first step to determine the effect of providing hand hygiene interventions on the level of knowledge and skills of school children.

## METHOD

The articles analyzed were obtained from search results across databases such as Google Scholar, Science Direct, Medline, and Pubmed using the keywords: delivery, education, health, hand hygiene, skills, students, elementary school. Articles reviewed are articles that have met the criteria based on the PICO, with a publication range from 2017-2021, which are written in Indonesian and English. The inclusion criteria must include the most effective and applicable methods in terms of time and tools to improve hand hygiene skills in elementary school students. The articles are then evaluated using a critical appraisal and in the form of a PRISMA guide. From the literature review, there are 20 (twenty) articles. The PRISMA flowchart used to determine the literature can be seen as follows:

**Figure 1.**
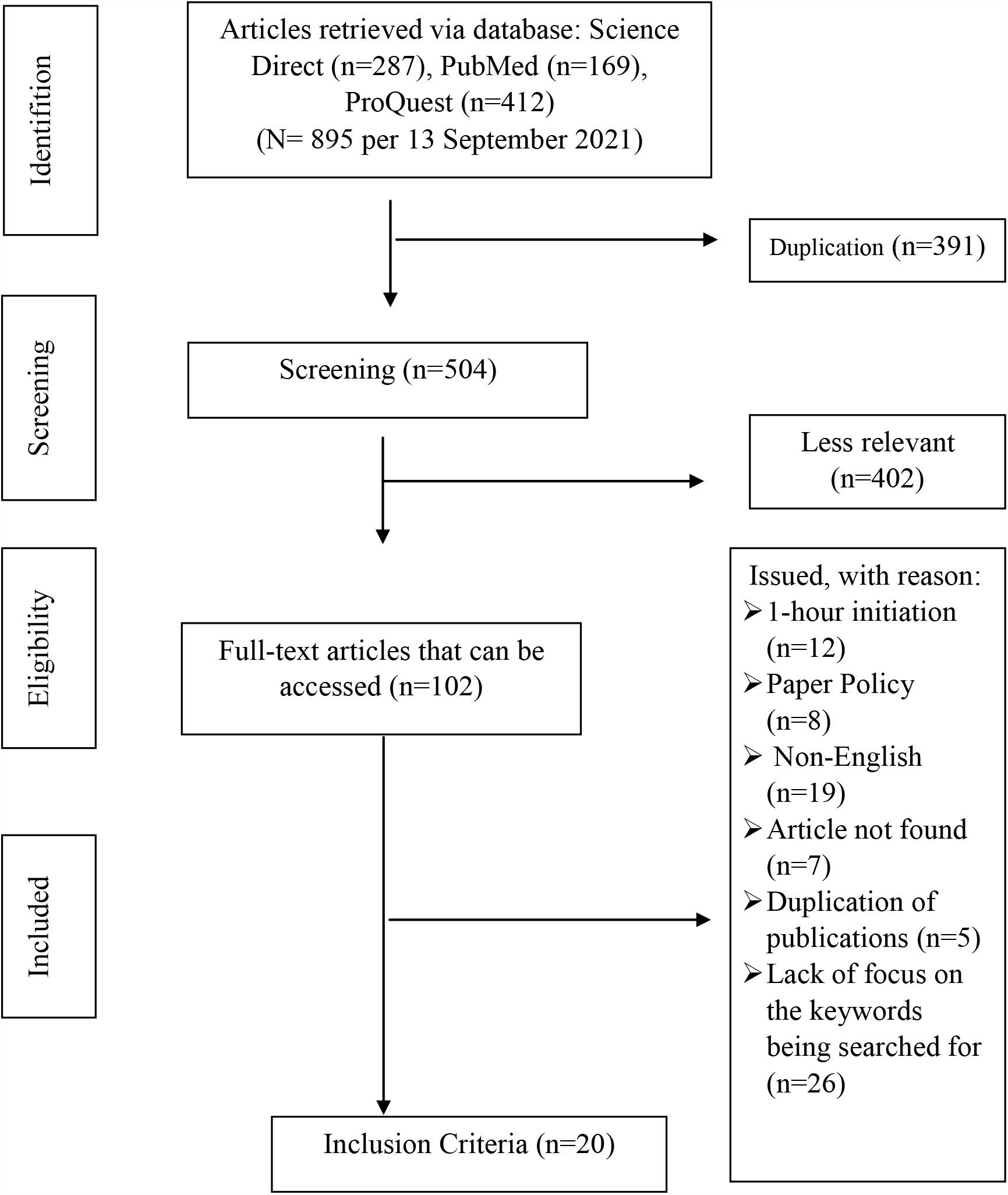
PRISMA Flowchart

## RESULT

**Table 1.** Summary of review results

| No | Authors and Titles | Purposes | Method (Design. Variables, Samples, and Instruments) | Result |
| --- | --- | --- | --- | --- |
| 1. | Ni Ketut Vera Parasyanti, et al; 2020<br>Health Education Handwashing with Soap using Video on the ability to wash hands in elementary school students | analyzed the effect of health education on hand washing using soap (CTPS) with video media on the ability to wash hands in third grade students | D: pre-experiment approach using *one group pretest posttest design*<br>S: 27 respondents<br>V: the ability to wash hands before and after watching the video.<br>I: checklist sheet and handwashing observation sheet that has been determined.<br>A: *Wilcoxon Signed Rank Test* (P value = 0,05). | After receiving health education about handwashing with soap using a video about the ability to wash hands, there was a significant change in the handwashing behavior of students in class III SDN 1 Berangbang Jembrana. |
| 2. | Arbianingsih, et al; 2018<br>Arbi Care Application Increases Preschool Children's Hand-Washing Self-Efficacy among Preschool Children | to test and determine the effectiveness of an Android mobile game application, namely ARBI CARE among preschool children as a means to prevent diarrhea and build self-efficacy in washing hands | D: Approach: *a pre-and post-test control group and time series design*<br>S: 60 respondents<br>V: self-efficacy, hand washing skills<br>I: Observation and questionnaire<br>A: GLMRM test | Android-based educational games can be an effective medium to increase handwashing self-efficacy in preschool children, thereby helping to prevent diarrhea. |
| 3. | Lorna Kwai, et al; 2020<br>Effectiveness of "Hand Hygiene | to determine the effectiveness of a 4-week series of educational sessions, | D: quasi-experimental<br>S: children aged 5-6 years in kindergarten (TK) | The results of this study have implications for school health educators and parents to promote hand hygiene (HH) |
|  | Fun Month" for Kindergarten Children: A Pilot Quasi-Experimental Study | covering the cognitive developmental stages of children, in increasing their knowledge and promoting hand hygiene practices. | V: Knowledge Level of Hand Hygiene Behavior<br>I: questionnaire and elements of the Hand Hygiene Education Program<br>A: Whitney U-test | practices among children at home and at the school level. |
| 4. | Ernestina A-M., et al; 2020.<br>Effectiveness of a hand hygiene program to reduce acute gastroenteritis at child care centers: A cluster-randomized trial | to assess the effectiveness of education and hand hygiene programs in child care centers (DCC) and homes on the incidence of acute gastroenteritis (AGE) in children attending DCC. | D: A randomized, controlled, open study<br>S: 911 respondents<br>V: Risk factors for acute gastroenteritis included in the stratified model<br>I: handwashing intervention.<br>A: A multilevel Poisson regression model | showed that a hand hygiene program that included hand sanitizer was most effective in the winter months with the highest incidence of AGE in children in DCCs. Furthermore, the greatest reductions in AGE episodes occurred in children who participated in a hand hygiene program including hand sanitizer and action education for DCC staff, parents, and children and were vaccinated for rotavirus. |
| 5 | Cheerful Hadem, et al; 2018<br>Effectiveness of Child To Child Approach on Knowledge and Practice Regarding Importance of Hand Washing Among Primary School Children | To evaluate the effectiveness of the child-to-child approach by comparing the pre and post-test values of knowledge and practice | D: Quasi-Experimental one group Pre-test Post-test<br>S: 60 respondents<br>V: knowledge, hand washing skills, child to child intervention<br>I: Questionnaire<br>A: test t-test | The child-to-child approach is an effective method for teaching health concepts such as hand washing among school children |
| 6. | Samantha L. Lange, et al; 2019<br>Effect of a simple intervention on hand hygiene-related diseases in preschools in South Africa: research protocol for an intervention study | The study aimed to determine whether HH interventions could reduce HH-related illnesses among 4-5-year-old preschool children and to improve HH practices in these children. | D: random distribution of control and experimental<br>S: 70 children aged 4-5 years<br>V: interactive learning, parental information, improvement of HH facilities<br>I: questionnaire and video<br>A: Poisson regression | HH behavior is self-reported and this may result in response bias, however, these results will be cross-checked with HH observations and swab hands |
| 7. | Daniel Eshetu, Tigist Kifle, et al ; 2020<br>Knowledge, Attitudes, and Practices of Hand Washing among Aderash Primary Schoolchildren in Yirgalem Town, Southern Ethiopia | To be able to find out about the knowledge, attitudes, and practices of handwashing in schoolchildren at SD Aderash in the city of Yirgalem. | D: A cross-sectional study<br>S: 279 respondents<br>V: Knowledge, attitude, hand washing practice<br>I: questionnaire<br>A: Epi-Info version 7 and exported to SPSS version 20 | There are >60% of school children have good knowledge and show a positive attitude towards hand washing, the correct handwashing practice is <40% |
| 8 | Julie Watson, et al; 2018<br>Child's play: Harnessing play and curiosity motives to improve child handwashing in a humanitarian setting | to determine the long-term behavioral and health impacts of such interventions when administered on a larger scale in a humanitarian context | D: controlled study design before and after (CBA)<br>S: 80 respondents<br>V: child characteristics, handwashing intervention, handwashing with soap.<br>I: questionnaire, cheat toy.<br>A: difference-in-<br>difference analysis via multilevel mixed-effects Poisson model. | Our results provide evidence that these ready-to-implement interventions can be effective in improving child handwashing practices in humanitarian settings while facilitating rapid implementation in the context of humanitarian emergencies. |
| 9 | Smitha M. M, et al; 2018<br>Effectiveness of Child-to-Child Approach on Practice of Hand Washing Among School Children in a Selected School at Mangalore | The effectiveness of the child-to-child approach in handwashing practice. | D: Quantitative evaluative research approach & pre-experiment with One group pre-test and post-test design<br>S: 75 respondents<br>V: knowledge, hand washing skills<br>I: questionnaire<br>A: and independent-t-test, chi-square, and fisher's exact test | Peer group training, child to child approach, and proper motivation are effective methods to teach children healthy habits such as hand washing and other common issues concerning children. |
| 10 | Sri Wahyuni, et al; 2017<br>Improving handwashing behavior with modeling techniques in the school-age group | to improve handwashing behavior with modeling techniques in elementary school-age children (6-12 years). | D: quasi-experimental with the non-equivalent group and before-after design<br>S: 38 intervention group and 38 control group<br>V: family characteristics of school children, the ability to wash hands before and after the modeling technique intervention<br>I: questionnaire, modeling technique intervention<br>A: independent t-test | The results showed that handwashing practice increased with modeling activity ($p=0.000$). Behavior modification with modeling techniques can be applied as an effort to improve handwashing behavior in school children which can be integrated into school nursing services |
| 11 | Afik Achsanti Saputri, Suryati; 2019<br>Effect of health education using audiovisual on knowledge of | to increase knowledge of washing hands with soap | D: pre-experiment with One-group pre-post test design<br>S: 44 respondents<br>V: Knowledge<br>I: Questionnaire<br>A: Wilcoxon Signed Ranks correlation | there is a change after being given health education using audiovisual on CTPS knowledge in grade IV children |
|  | handwashing with soap (CTPS) in fourth-grade children at MI Jamilurrahman Bantul |  | test |  |
| 12 | Mayuri A. Mane, et al; 2017<br>A Study to Assess the Effectiveness of Hand Hygiene Technique among School Children in Maharashtra, India | to assess the effectiveness of hand hygiene techniques demonstration of hand hygiene practices in elementary school children. | D: one group pre-test, post-test design<br>S: 60 respondents<br>V: knowledge, hand washing skills<br>I: SOP for washing hands<br>A: Descriptive and inferential statistics by considering the objectives | demonstration of hand hygiene techniques on hand hygiene is considered effective in improving hand hygiene practices in elementary school children so that the hand hygiene technique steps are easy to understand. |
| 13 | Octa, A; 2019<br>The relationship between knowledge and attitudes towards handwashing behavior in the Pegirian village community | Identifying the relationship between knowledge and attitudes towards hand washing in RW III Pegirian Village | D: quantitative with cross-sectional design<br>S: 84 respondents<br>V: knowledge, attitude<br>I: Questionnaire<br>A: Spearman correlation test | The existence and strength of the relationship between knowledge and attitudes towards hand washing is positive |
| 14 | Agus Erwin Ashari, et al; 2020<br>Improving Knowledge, Attitudes, and Practices of Handwashing with Soap in Class V Elementary School Children | knowing and analyzing the relationship between Handwashing with Soap (CTPS) and increasing knowledge, attitudes, and practices of hand hygiene using soap at the | D: non-equivalent pre-post test control group design<br>S: 61 respondents<br>V: knowledge, attitude, hand washing practice<br>I: Questionnaire<br>A: Wilcoxon test | The results showed that there were no significant differences in knowledge, attitudes, and practices before and after the test in the control group, and no significant differences in knowledge, attitudes, and practices before and after CTPS training in the intervention group. There was a |
|  | through Handwashing with Soap Exercise. | age of elementary school children in Mamuju District |  | difference in knowledge between the intervention group and the control group, but no difference in attitude or practice. |
| 15 | Padila, et al; 2020<br>Learning to wash hands in seven steps through the demonstration method for early childhood | To evaluate the effectiveness of the demonstration method for hand washing skills at 7 stages of preschool children in Aisyiyah 1 Kindergarten, Bengkulu City | D: pre-experimental design with one group pretest-posttest<br>S: 30 respondents<br>V: The ability to wash hands before and after the demonstration<br>I: SOP for washing hands<br>A: Wilcoxon test | Demonstration intervention is very effective in increasing the ability to wash hands by 7 steps in early childhood. |
| 16 | Yulianto, et al; 2018<br>The effectiveness of media ( video, image, and song) to Handwashing behavior in 1<sup>ST</sup> – 3<sup>RD</sup> graders of SDIT permata Mulia Mojokerto | to find out the effectiveness of the video, image, and song method on handwashing behavior | D: Pre-experiment design with pre-post test design<br>S: 90 respondents<br>V: handwashing behavior, hand washing skills intervention<br>I: videos, pictures, songs.<br>A: compare mean scores | The song method is more effective than the simulation method in improving the ability to wash hands |
| 17 | Tri Nugroho, et al; 2020<br>The Effect of Health Education Using Audio-Visual Media on Knowledge of Hand Washing | to be able to determine the effect of health education with audiovisual media on knowledge of handwashing with soap in grade 2 | D: pre-experiment with one group pretest-posttest approach<br>S: 46 respondents<br>V: Knowledge before and after being given audio-visual media washing hands | There has been a change since the introduction of health education through audiovisual interventions given, to increase knowledge about handwashing with soap. |
|  | with Soap in Grade 2 Elementary School Children | elementary school children | I: questionnaire<br>A: Wilcoxon Signed Ranks Test |  |
| 18 | Rina Mariyana, et al; 2021<br>The application of the animated video for washing hands with soap on the response to children at a special school in Bukittinggi | To introduce an animated video intervention for washing hands with soap as a response to reactions to children from the Bukittinggi Special School | D: sequential exploratory<br>S: 34 respondents<br>V: knowledge, attitude, action<br>I: Questionnaire, video implementation<br>A: chi-square test | Hand washing video intervention must be carried out on children, and if the video is played continuously then the child's needs and abilities can increase the effectiveness in washing hands |
| 19 | Yesica Devis, et al; 2018<br>Lecture Methods and Effective Group Discussions Increase Students' Knowledge About Hand Washing With Soap | Seeing the effectiveness of the lecture method and group discussion in increasing the knowledge of fourth-grade students about hand soap at SDN 044 Pekanbaru | D: quasi-experiment using a one group pre-test post-test approach<br>S: 68 respondents<br>V: knowledge<br>I: questionnaire<br>A: independent T-test and Mann-Whitney test | There is a difference in the average knowledge score after the intervention using either the lecture method or the group discussion method. However, the group discussion method is better. |
| 20 | Balwani C.M, et al; 2019<br>Effect of a school-based hand hygiene program for Malawian children: A cluster randomized controlled trial | to evaluate the impact of a school-based hand hygiene program on children's handwashing adherence and school absenteeism in South Africa | D: 2-arm cluster RCT.<br>S: 6 elementary schools with 375 children<br>V: Hand washing knowledge, hand washing skill mastery, hand hygiene, and absenteeism rate increase under 5 simplified steps<br>I: questionnaire, hand washing soap | The impact of the school-based hand hygiene program is positive. It can be used in both the planning and development of hand hygiene protocols to increase schoolchildren's handwashing compliance rates and to reduce school absenteeism in developing countries. |
|  |  |  | A: SPSS program (software) versions 23.0; IBM Corp, Armonk, NY) |  |

## DISCUSSION

Based on the results of a systematic literature review on the effect of providing hand hygiene interventions on handwashing skills in elementary school students, namely as follows:

When searching for hand hygiene articles in this study, it was found that there were 10 studies with quasi-experimental and pre-experimental designs. This study found that there was an impact of hand hygiene interventions using different methods on the level of knowledge and skills indicated by a significant improvement in hand hygiene practices.

The selection of methods and media also determines the effectiveness of the intervention. As in the research conducted by (Ni Ketut Vera P, 2020) which showed that there was an impact of health education on handwashing with soap using video media on the ability to wash hands for class III SDN 1 Berangbang Jembrana. This is also in line with research by (Afik Achsanti Saputri, Suryati; 2019) on the effect of audiovisual health education on CTPS knowledge for Class IV MI Jamilurrahman Bantul children. In addition, different simulation interventions were carried out by (Yulianto, et al; 2018) with the song method which was more effective than the simulation method in improving the ability to wash hands. Meanwhile, in the study (Agus Erwin Ashari, et al; 2020) the results showed that there were no significant differences in knowledge, attitudes, and practices between the pre-test and post-test in the control group. In the intervention group, there was no significant difference in knowledge, attitude, or practice before and after CTPS exercise. There is a difference in knowledge between the intervention group and the control group, but there is no difference in attitude and practice

When deciding on methods and media, the effectiveness of the intervention is also needed. Several studies, such as research by Arbianingsih (2018), show that the use of intervention strategies in the form of games (android-based educational games) can be an effective medium to increase handwashing self-efficacy in preschool children, thereby helping prevent diarrhea. Even in research (Rina Mariyana, et al; 2021) to activate children’s ability to wash their hands, it can be taught through animated videos that are played repeatedly.

Based on the evaluation of the literature that has been obtained, it is known that there is an effect of various hand hygiene interventions on the knowledge and skills of elementary school students through the lecture and role-playing methods combined with audiovisual media. This is because, according to the survey above, the average skill level reaches scores were quite high, and in most studies, the health education intervention improved after washing hands with audiovisual media such as lectures, demonstrations, and role-playing. The increase was also higher in the intervention group than in the control group.

## CONCLUSION

In the collection of literature contained in this study, it can be concluded that there is a significant increase in handwashing behavior in students who are intervened with health education methods. demonstrations and audiovisuals. In addition, the selection of attractive media also affects the effectiveness of an intervention. Although some of the literature studies obtained have intervention strategies to improve other hand hygiene skills, lecture and role-playing methods and the use of audiovisual intervention strategies have been shown to make a significant difference after the intervention increased.

## Data Availability

Conflict of Interest Disclosure Form
All authors need to complete and submit this form when submitting a manuscript. Disclosures and signatures from all authors on one form is preferred.
Article title:
THE EFFECT OF HAND HYGIENE INTERVENTION ON THE KNOWLEDGE AND SKILLS LEVEL OF SCHOOL CHILDREN :
A LITERATURE REVIEW
Please note that a conflict of interest statement is published with each paper and must be inserted in your text document right before the reference list.
√ I/we certify that there is no actual or potential conflict of interest in relation to this article.
(Please print names)
(1st author): Nurlela Mufida
Signature:
Date: 4st January, 2022
(2nd author): MOSES GLORINO RUMAMBO PANDIN
Signature:
Date: 4st January, 2022
Corresponding Author: Name:
E-mail:

http://www.medrxiv.org

## REFERENCES

1. Afik Achsanti Saputri, Suryati. (2019). Pengaruh pendidikan kesehatan menggunakan audiovisual Terhadap Pengetahuan Cuci Tangan Pakai Sabun (CTPS) Pada Anak Kelas IV Di MI Jamilurrahman Bantul; Jurnal Medika Respati Vol. 14 (3) Juli 2019. DOI: https://doi.org/10.35842/mr.v14i3.231

2. Agus Erwin Ashari, dkk. (2020). Peningkatan Pengetahuan, Sikap Dan Praktik Cuci Tangan Pakai Sabun Pada Anak Kelas V Sekolah Dasar Melalui Senam Cuci Tangan Pakai Sabun ; Jurnal Ilmiah Permas: Jurnal Ilmiah STIKES Kendal Vol 10 (1), Hal 11 - 18, Januari 2020. DOI: https://doi.org/10.32583/pskm.v10i1.635

3. Arbianingsih, et al. (2018)Arbi Care Application Increases Preschool Children’s Hand-Washing Self–Efficacy among Preschool Children; Enferm Clin. 2018;28(Suppl 1 Part A):27–30. https://doi.org/10.1016/S1130-8621(18)30031-7

4. Audria Octa Anggraini Widi Lestari. (2019). Hubungan pengetahuan dan sikap terhadap perilaku cuci Tangan pada masyarakat kelurahan pegirian; Jurnal Promkes Vol. 7 (1) hal ; 23-34. doi: 10.20473/jpk.V7.I1.2019.1-11

5. Balwani Chingatichifwe Mbakaya, et al. (2019).Effect of a school-based hand hygiene program for Malawian children: A cluster randomized controlled trial; American Journal of Infection Control 00 (2019) 1−5. DOI: 10.1016/j.ajic.2019.06.009

6. Cheerful Hadem, et al. (2018). Effectiveness of Child To Child Approach on Knowledge and Practice Regarding Importance of Hand Washing Among Primary School Children; RGUHS Journal of Nursing Sciences, June 2018 / Vol-8 / Issue-1.

7. Daniel Eshetu, Tigist Kifle, et al. (2020). Knowledge, Attitudes, and Practices of HandWashing among Aderash Primary Schoolchildren in Yirgalem Town, Southern Ethiopia; Journal of Multidisciplinary Healthcare 2020:13 759–768. DOI: 10.2147/JMDH.S257034

8. Ernestina A.M, et al. (2020). Effectiveness of a hand hygiene program to reduce acute gastroenteritis at child care centers: A cluster-randomized trial; American Journal of Infection Control 000 (2020) 1−7. https://doi.org/10.1016/j.ajic.2020.03.011.

9. Julie Watson, et al. (2018). Child’s play: Harnessing play and curiosity motives to improve child handwashing in a humanitarian setting; International Journal of Hygiene and Environmental Health.https://doi.org/10.1016/j.ijheh.2018.09.002

10. Lorna Kwai, et al (2020). Effectiveness of “Hand Hygiene Fun Month” for Kindergarten Children: A Pilot Quasi-Experimental Study; Int. J. Environ. Res. Public Health 2020, 17, 7264; doi:10.3390/ijerph17197264

11. Mayuri A. Mane, et al. (2017) A Study to Assess the Effectiveness of Hand Hygiene Technique among School Children in Maharashtra, India; Asian Journal of Pharmaceutical Research and Health Care, Vol 9(4), 174–179, 2017. DOI: 10.18311/ajprhc/2017/15834

12. Ni Ketut Vera Parasyanti, et al (2020) Pendidikan Kesehatan Cuci Tangan Pakai Sabun dengan Video Terhadap kemampuan cuci tangan pada siswa SD; Jurnal Akademika Baiturrahim Jambi, Vol. 9 (1) hal:122-130. Doi : 10.36565/jab.v9i1.197

13. Padila, dkk. (2020). Pembelajaran cuci tangan tujuh langkah melalui Metode demonstrasi pada anak usia dini ; Journal of Telenursing (JOTING)Volume 2, Nomor 2, Desember 2020. DOI: https://doi.org/10.31539/joting.v2i2.1395

14. Rina Mariyana, dkk. (2021). Penerapan vidio animasi cuci tangan pakai sabun Terhadap respon pada anak Di sekolah luar biasa bukittinggi ; Jurnal Endurance : Kajian Ilmiah Problema Kesehatan Vol 6(1) Februari 2021 (38-45)

15. Smitha Mariam Mathew, et al (2018). Effectiveness of Child To Child Approach on Practice of Hand Washing Among School Children in a Selected School at Mangalore; Journal of Health and Allied Sciences NU 2018; 08(01): 15–21. DOI: 10.1055/s-0040-1708739

16. Sri wahyuni, dkk. (2017). Peningkatan perilaku mencuci tangan dengan Teknik modeling pada kelompok anak usia Sekolah; The Indonesian Journal Of Health Science, Vol. 8 (2) Juni 2017. DOI: https://doi.org/10.32528/the.v8i2.868

17. Tri Nugroho, dkk. (2020). Pengaruh Pendidikan Kesehatan Dengan Media Audio Visual Terhadap Pengetahuan Cuci Tangan Pakai Sabun Pada Anak Sd Kelas 2 ; Healthy Journal Vol. VIII No. 1, Maret 2020.

18. Yesica devis, dkk. (2017). Metode Ceramah Dan Diskusi Kelompok Efektif Meningkatkan Pengetahuan Siswa Tentang Cuci Tangan Pakai Sabun ; Jurnal Kesehatan Komunitas 2017;3(4):159–163. https://doi.org/10.25311/keskom.Vol3.Iss4.205

19. Yulianto, et al (2018). The effectiveness of media (video, image, and song) to Handwashing behavior in 1^ST^ – 3^RD^ graders of SDIT Permata Mulia Mojokerto; International Journal Of Nursing and Midwifery Science (IJNMS), Volume2, Issue 2, August 2018. DOI: https://doi.org/10.29082/IJNMS/2018/Vol2/Iss02/133

20. Samantha Louise Lange, et al. (2019). Effect of a simple intervention on hand hygiene-related diseases in preschools in South Africa: research protocol for an intervention study; BMJ Open 2019;9:e030656. doi:10.1136/bmjopen-2019-030656

